# Effect of Hyperbaric Oxygen After Moderate-Intensity Exercise on Fatigue: A Single-blind Crossover Randomized Trial

**DOI:** 10.1101/2025.01.19.25320809

**Authors:** Kazuyoshi Yagishita, Junya Aizawa, Shunske Ohji, Takashi Hoshino, Takuya Oyaizu

**Author notes:** Corresponding Author (KY).

## Abstract

**Objectives:** Although insufficient delivery of oxygen might be a factor in physical and perceived fatigue, the relationship between exposure to hyperbaric oxygen (HBO) and recovery from perceived fatigue remains unexplained. The purpose of this study was to investigate the effects of exposure to HBO after long-duration, medium-intensity training on recovery from perceived fatigue in a single-blind study.

**Methods:** Fatigue was induced in nine male university students (mean age: 21.3 years) using an ergometer exercise bike at a moderate intensity of 75% of their maximum heart rate for 60 min. Post-workout, subjects randomly received an intervention comprising exposure to HBO or an air placebo in a single-blind experimental trial. Blood tests were conducted, and perceived fatigue was evaluated by using visual analog scales (VAS) at five time points. One week later, a crossover trial was conducted.

**Results:** For the HBO group, pre- to post-intervention mean VAS scores for whole-body fatigue significantly improved from 48.4 to 28.7 (p<0.001). However, in the placebo group, the improvement was not statistically significant. Hematological evaluations, including C-reactive protein level, white blood cell count, creatine kinase level, lactic acid level, T-cell count, CD4/CD8 ratio, and natural killer cell count, did not show any significant between-group differences except only BUN and free fatty acids levels 1.5 h after intervention and Mg levels immediately after intervention. There were also no significant differences in the results of other hematological assessments.

**Conclusions:** The results of this study showed that perceived fatigue evaluated by VAS scores significantly improved in the HBO group in a blinded trial. However, this was not objectively supported by blood test results. HBO may have effects on recovery from perceived fatigue following long-duration, moderate-intensity exercise. However, the results of this study could not determine the efficacy of HBO on exercise-induced fatigue.

## Introduction

Hyperbaric oxygen (HBO) treatment is defined as an intervention in which an individual breathes near 100% oxygen intermittently while inside a hyperbaric chamber that is pressurized to greater than the sea level pressure (1 atmosphere absolute, ATA). For clinical purposes, the pressure must be equal to or exceed 1.4 ATA while breathing near 100% oxygen (1). HBO can increase serum dissolved oxygen levels in proportion to oxygen partial pressure and transport dissolved oxygen to ischemic tissue. HBO is an established treatment for several conditions such as decompression illness, carbon monoxide poisoning, acute arterial disturbance, peripheral ischemic disease, compartment syndrome, and delayed radiation injury.

The effects of HBO treatment on soft tissue injuries during sports activities, including sprains (2, 3), ligament injuries (4–7), contusions, and muscle strains (8–13), have been widely reported. HBO is an effective treatment for edema reduction and improves local perfusion and oxygenation of the injured tissues (3, 13–15).

However, there is disagreement regarding the effectiveness of HBO treatment in promoting recovery from exercise-induced fatigue. Studies suggest that it has a positive effect; a study has shown that HBO treatment for delayed onset muscle soreness (DOMS) resulting from eccentric training of the quadriceps promoted eccentric torque recovery (16). More recently, a study of the effects of HBO treatment on muscle fatigue after repeated plantar flexion of the ankle joint found that HBO treatment reduced the decline in muscle force production (17). Further, in a study of Brazilian jiu-jitsu athletes, although no HBO intervention-related differences were found in the post-training blood tests, a significant difference in scores on a perceived recovery scale was found, suggesting its effect on self-perceived recovery (18). Finally, an insufficient delivery of oxygen has also been shown to be a factor in the development of central and peripheral fatigue (19).

In contrast, studies have also found that HBO treatment results in no significant difference in recovery from DOMS caused by eccentric workouts of the elbow or knee flexor muscle groups (20–22). Further, a Cochrane Review of research on the effectiveness of HBO for DOMS and closed soft tissue injury treatment found that evidence was insufficient to establish its superiority over other treatments (12). Therefore, studies that are carefully designed for validity would be needed to establish the effectiveness of HBO treatment for the treatment of fatigue. Thus, this study was designed as a high-quality single-blind, crossover randomized study with detailed multiple-biomarker items.

We hypothesized that HBO would reduce fatigue after long-duration, moderate-intensity exercise. The purpose of this study was to test the effectiveness of treating healthy male university students who regularly exercised with HBO for fatigue after exercise with a single-blind and crossover randomized trial.

## Materials and Methods

### Trial design

The study used a crossover design in which all subjects received both the HBO intervention and air placebo following a training session with an interval of 1 week. For the first trial, they were randomly assigned to either the HBO or air placebo group. Randomization was allocated by the even or odd number of dice (KY), and participants were registered (TO). Only the researchers knew the treatment that each subject was receiving, making it a single-blind study. The validity of the study design was ensured using a single-blind crossover randomized trial and tests for physical and mental effects using blood tests and self-rated evaluation scales.

The protocol for this study was approved by the institutional review board of Tokyo Medical and Dental University (TMDU No. 2000-901). The application period for this research is from January 4th to March 31st, 2012. The trial registry is the ISRCTN registry, and the registration number is ISRCTN81080077.

### Subjects

The subjects were healthy male university students who exercised regularly. To be eligible, students needed to be at least ≥20 and <30 years of age at the time they gave their informed consent. Receipt of written consent was considered as an indication that the student had voluntarily agreed to participate after having received and understood a thorough explanation of the study, and written informed consent was obtained from all subjects prior to participation. Eligible students were eliminated from the sample if they had difficulty relieving pressure in their ears (which precludes or makes HBO treatment difficult), claustrophobia, congenital pulmonary cysts, asthma, a history of pneumothorax, or heart disease or if they had experienced heart palpitations, precordial pain, or tachycardia during the previous year. The sample size was calculated to be 15 people based on an alpha error of 0.05, a detection rate of 0.8, and an effect size of 0.5. In the protocol for the ethics review, there were 15 people in each of the HBO, oxygen, and air groups. However, the oxygen group was not implemented, and the study was changed to a crossover study of the HBO group and the air group. In reality, 9 students were recruited. None of the eligible students met any of the exclusion criteria. The date were collected at Tokyo Medical and Dental University Hospital.

### Exercise protocol

Fatigue was induced by having the subjects ride an ergometer exercise bike (Aerobike 75XL, COMBI Wellness Corp., Tokyo, Japan) at a medium intensity of 75% of their maximum heart rate, with a target pulse rate of 150 per minute for 60 min.

### Intervention and placebo

The HBO chamber in our university hospital was used; this multi-place HBO chamber can hold a maximum capacity of 16 individuals (NHC-412-A, Nakamura Tekko-Sho Corp., Tokyo, Japan). The HBO treatment protocol included 60 min of inhaling pure oxygen using a mask at pressures up to 2.5 ATA with two 5-min breaks to breathe air, 15 min for compression and 15 min for decompression for a total of 100 min (Fig. 1). This treatment protocol is the standard for HBO treatment and is used globally.

**Fig. 1.**
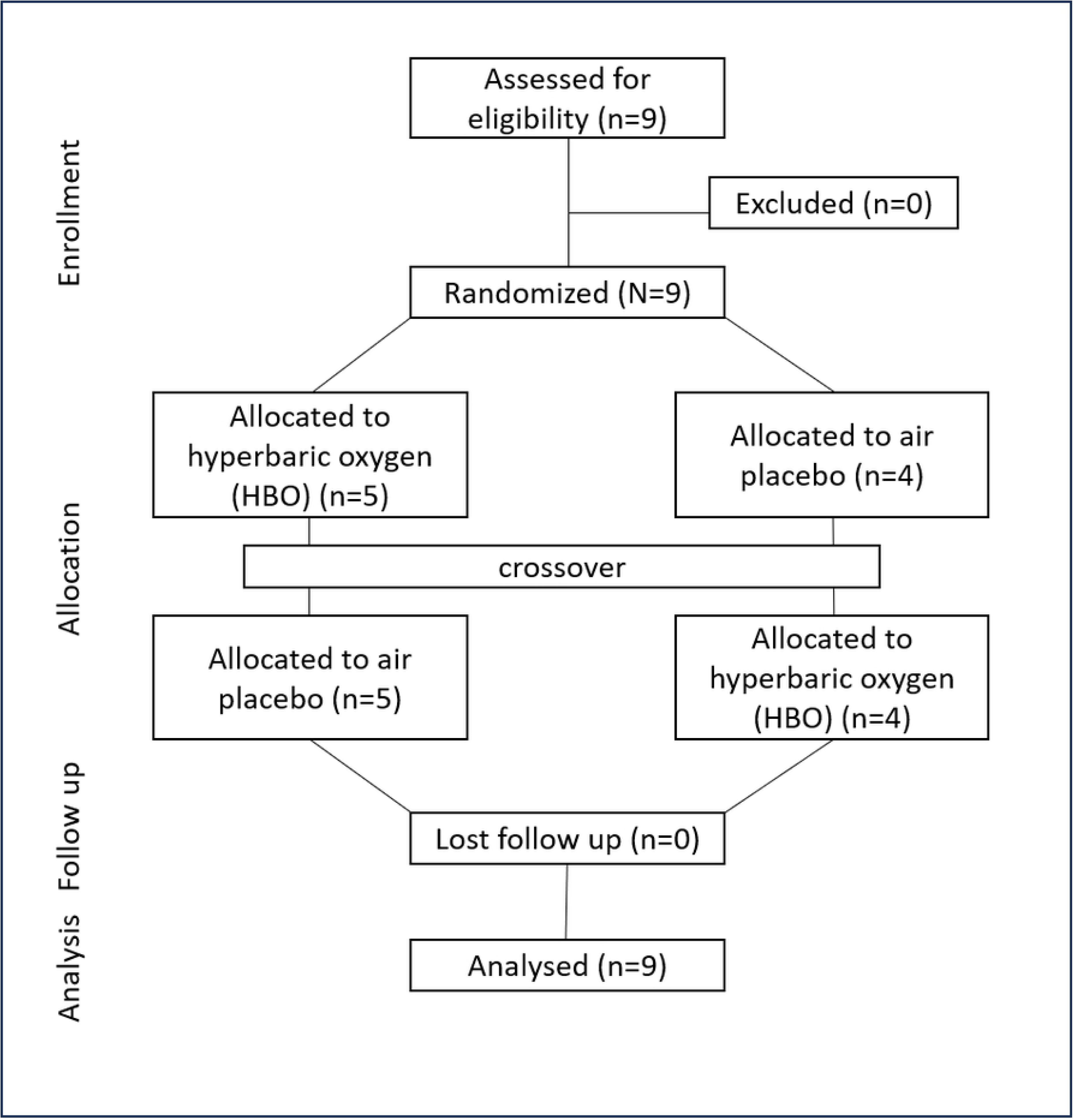
A flowchart of the experimental design.

For the protocol for air placebo, we chose a pressure and time duration safe enough to prevent decompression sickness. The intervention consisted of 80 min of breathing air at pressures up to 1.2 ATA, with 10 min for compression and 10 min for decompression, for a total of 100 min (Fig. 2) (23).

**Fig. 2.**
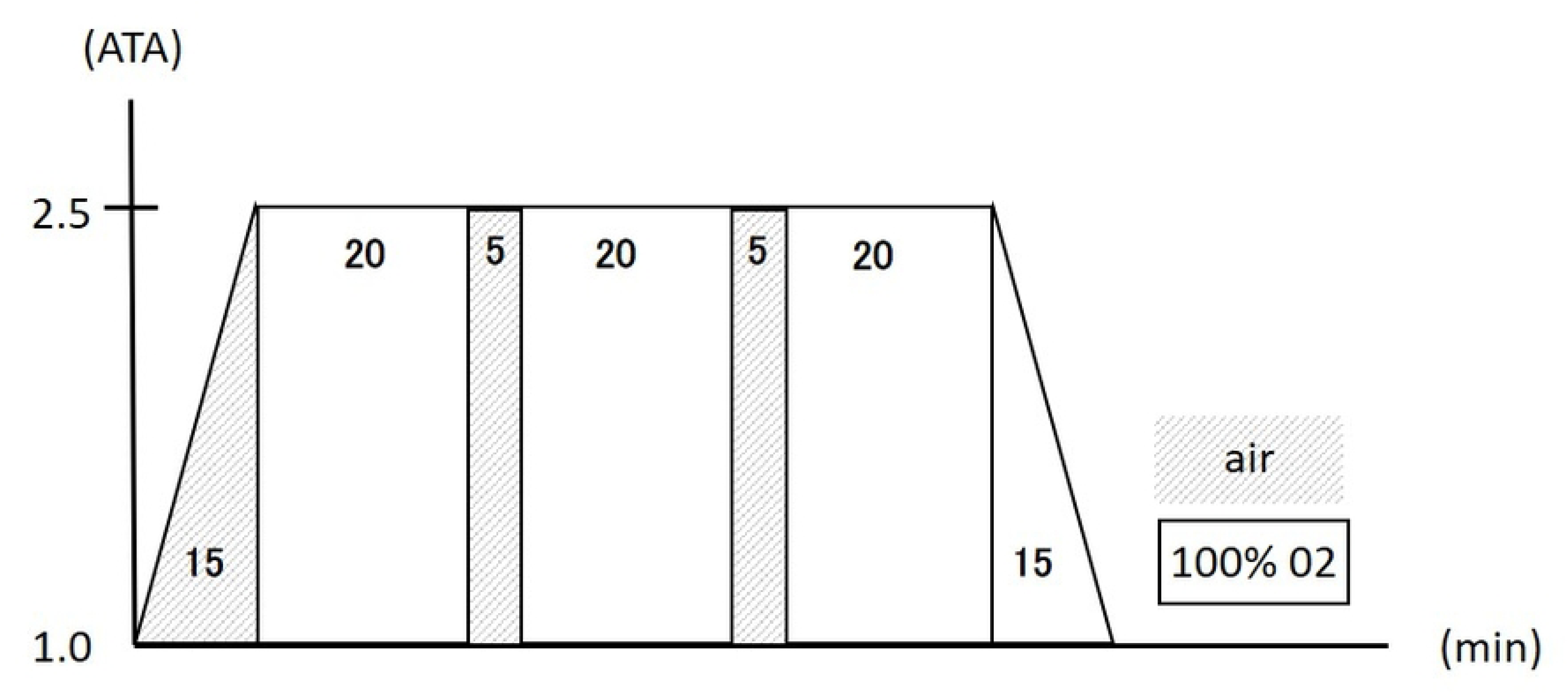
Protocol of hyperbaric oxygen intervention.

To ensure that subjects could not tell whether they were being treated with HBO or air, the pressure gages in the treatment room were hidden from their view and covered with a piece of cloth, and the tubes delivering oxygen and air were also kept out of sight.

In addition, to investigate the reliability of this single-blind study, after subjects received the intervention, they were enquired whether they thought they had received the HBO or air placebo. The question was asked to the nine subjects a total of 18 times. Three responses from three subjects were invalid. The remaining 15 valid responses were used to determine the correct response rates.

### Measures

#### 1) Visual analog scales (VAS)

VAS was used to measure perceived fatigue in their whole-body and leg. VAS score consisted of 100 points at full mark, with the worst condition designated as 100 points and no complaint as 0 point.

#### 2) Blood tests

Complete blood count: white blood cell (WBC), WBC differential, red blood cells, hemoglobin, hematocrit, and platelets.

Biochemical tests: C-reactive protein, erythrocyte sedimentation rate, aspartate transaminase, alanine transaminase, zinc tolerance test, alkaline phosphatase, lactate dehydrogenase (LDH), γ-glutamyl transpeptidase, cholinesterase, creatinine kinase (CK), total protein, creatinine, blood urea nitrogen (BUN), uric acid, triglycerides, total cholesterol, amylase, glucose, free fatty acids, albumin, lactate, bilirubin, potassium (K), calcium, magnesium (Mg), and myoglobin.

Immune system tests: interleukin-1β (IL-1β), IL-6, tumor necrosis factor-α (TNF-α), CD4, CD8, and natural killer (NK) cell count.

### Measurement timing

Blood tests and VAS scores for fatigue were measured pre-exercise (test 1), post-exercise (test 2), post-intervention (test 3), 1.5 h post-intervention (test 4), and 24 hours post-intervention (test 5). The first and second trials (crossover trial) were conducted 1 week apart (Fig. 3).

**Fig. 3.**
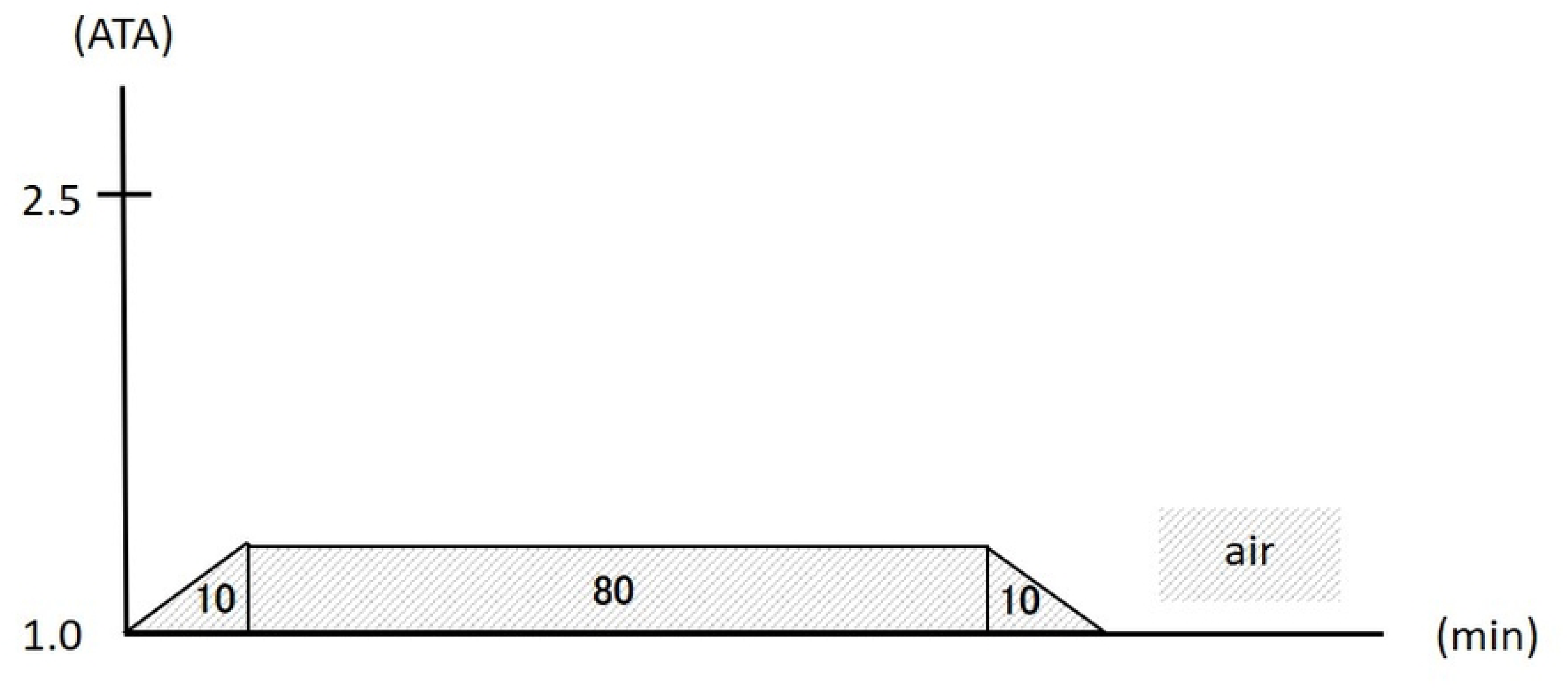
Protocol of air placebo intervention.

In the initial trial, five subjects received HBO treatment and four received placebo. Subsequently, in the crossover trial, four subjects received HBO treatment and five received placebo.

### Statistical analysis

Data were expressed as mean ± standard deviation and analyzed using SPSS version 19.0 (SPSS Inc., Chicago, IL, USA). The VAS scores or laboratory data between the HBO and air groups were evaluated using two-way analysis of variance followed by Bonferroni post-hoc test, and data between the tests were evaluated using paired t-test. p<0.05 was considered statistically significant.

## Results

### Baseline

All nine subjects completed the experiment. Their mean age was 21.3 ± 1.2 years (20‒23 years), mean height was 172.2 ± 6.2 cm, and mean weight was 82.3 ± 15.7 kg.

### Reliability of single-blinded study

Of the 8 subjects who received the HBO intervention, 3 answered correctly and 5 answered incorrectly as “air”. Of the 7 subjects who received the air placebo, 5 answered correctly and 2 answered incorrectly as “HBO”. The accuracy rate was 53.3%, indicating the high reliability of this blinded trial (Fig. 4).

**Fig. 4.**
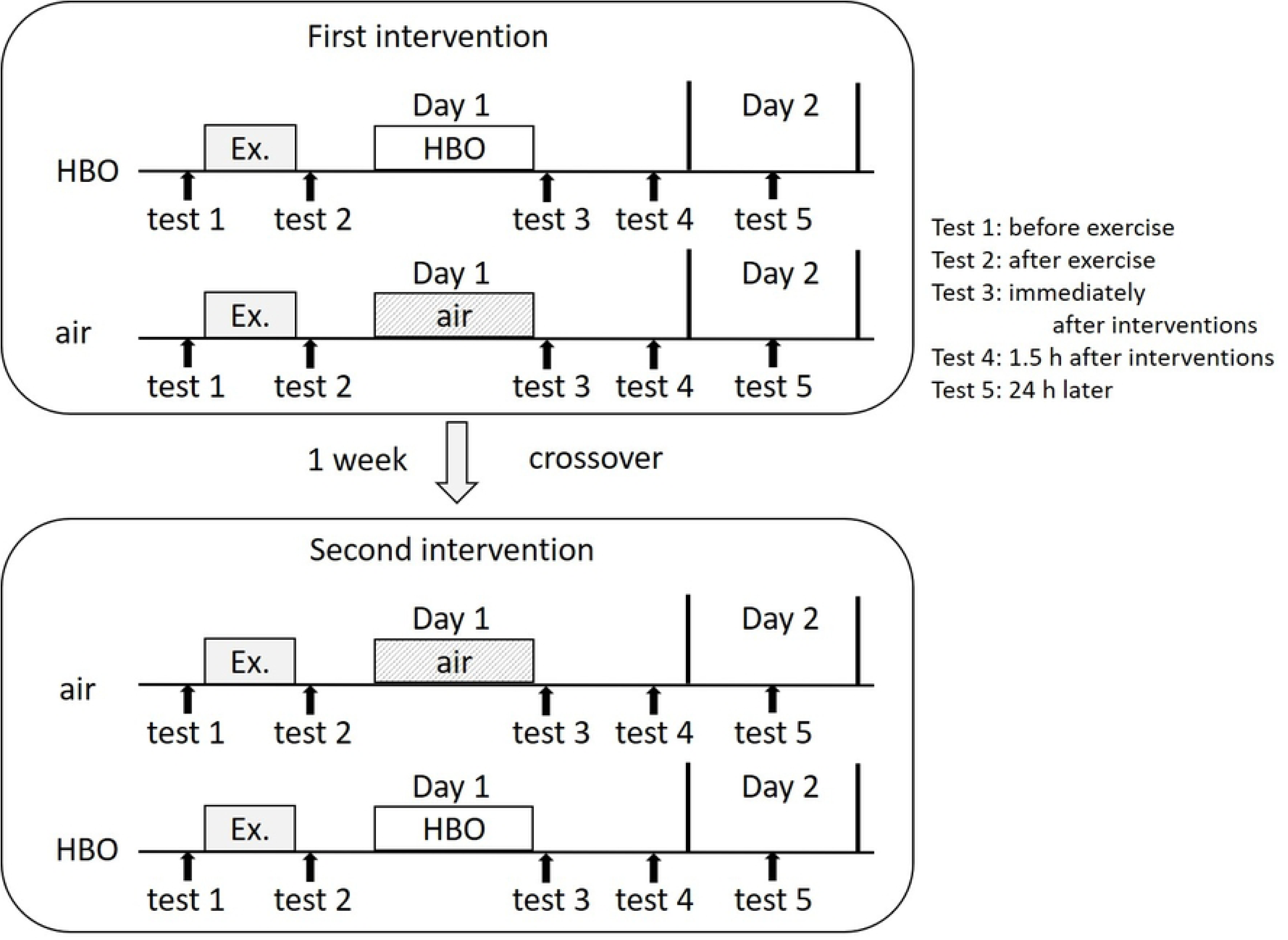
Measurement timing in crossover trial: test 1, before exercise; test 2, after exercise; test 3, immediately after interventions; test 4, 1.5 h after interventions; and test 5, 24 h later.

### Side Effects

No side effects were observed.

### VAS evaluation

Regarding the VAS scores (Fig. 5-a, -b), in the HBO group, the mean scores for whole-body fatigue at pre- to post-intervention (tests 2 and 3, respectively) significantly decreased from 48.4 ± 17.3 to 28.8 ± 18.6 (p<0.001), whereas in the air group, the decrease was from 43.2 ± 21.2 to 37.9 ± 24.1, which was not statistically significant (Fig. 5-a). There were no significant differences in data between the HBO and air groups. For leg fatigue, in the HBO group, the mean scores at pre- to post-intervention (tests 2 and 3, respectively) significantly decreased from 49.9±18.4 to 26.1±16.9 (p<0.001), and in the air group, these significantly decreased from 54.7±17.1 to 43.6±24.0 (p<0.05) (Fig. 5-b). Moreover, a significant difference in test 3 data was found between the HBO and air groups (p<0.05).

**Fig. 5.**
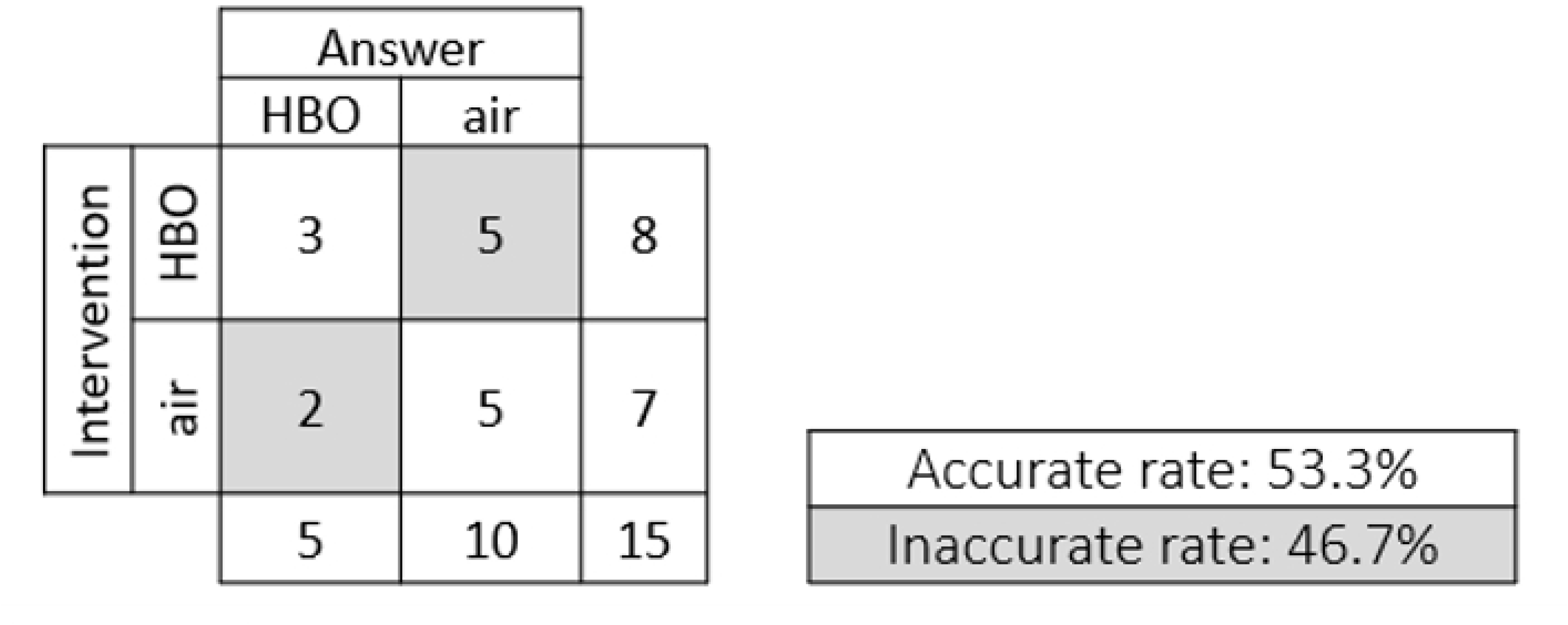
Assessment of reliability of the blinded intervention.

### Blood test

Comparative analysis of blood test (Fig. 6) results between the two groups showed significantly higher BUN and free fatty acids levels in test 4 (p<0.05) and Mg levels in test 3 (p<0.05) in the HBO group. No other significant differences in any of the results including lactate were found.

**Fig. 6.**
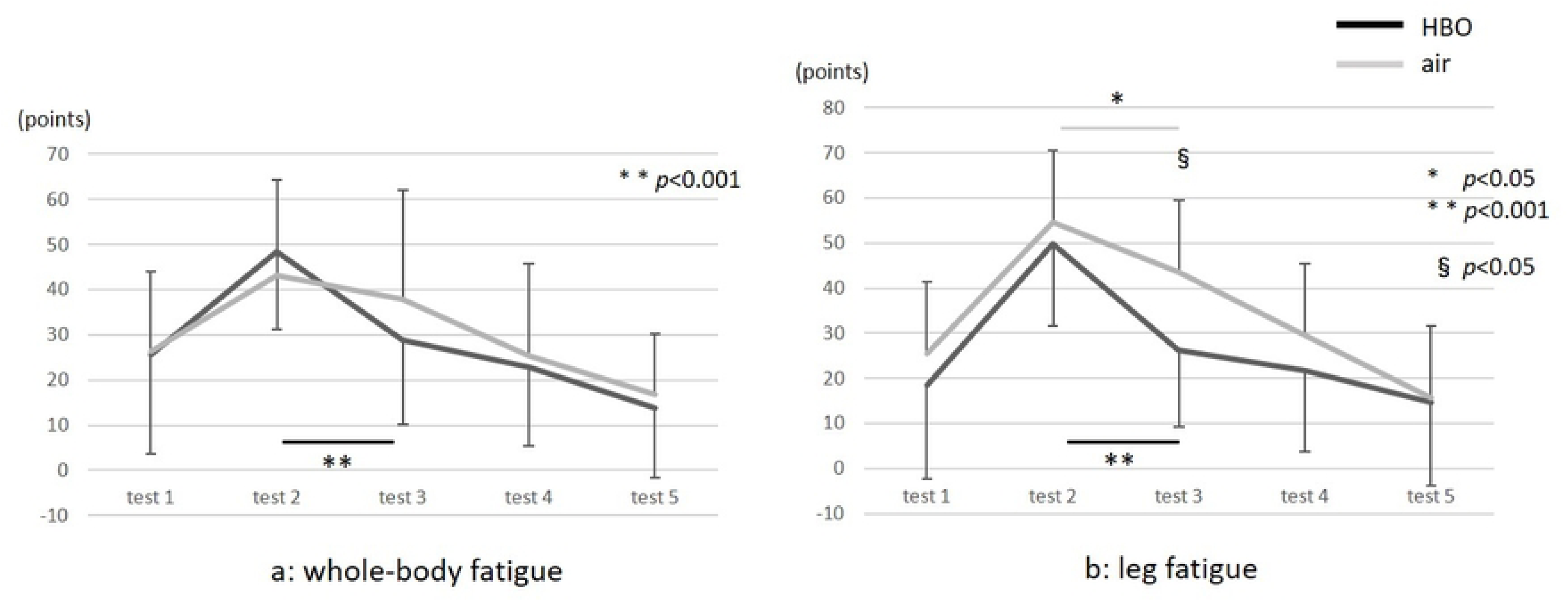
-a, b. Results of the visual analog scale of whole-body and leg fatigue. * and ** indicate the comparison between the tests, and § indicates the comparison between the groups.

**Fig. 7.**
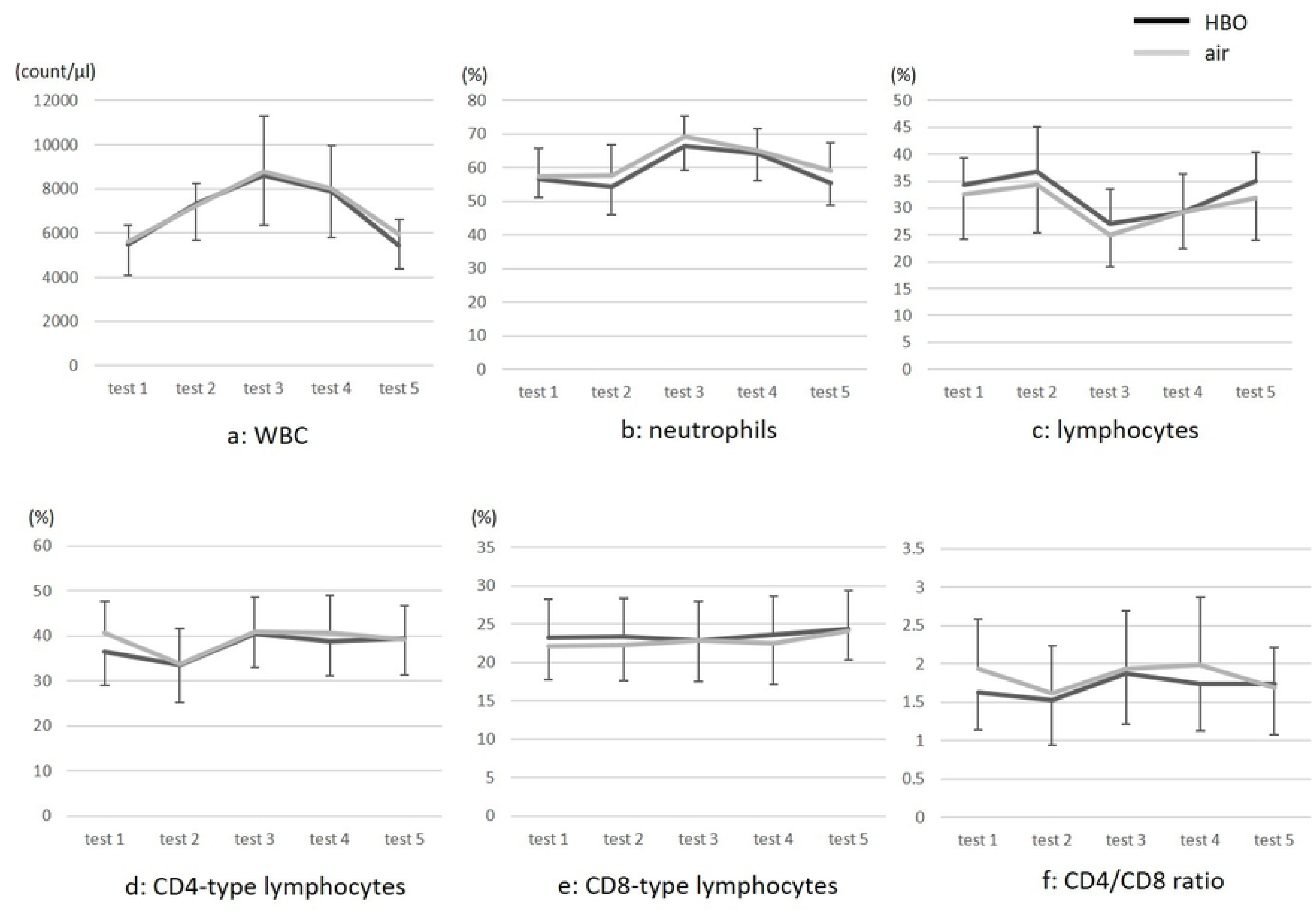
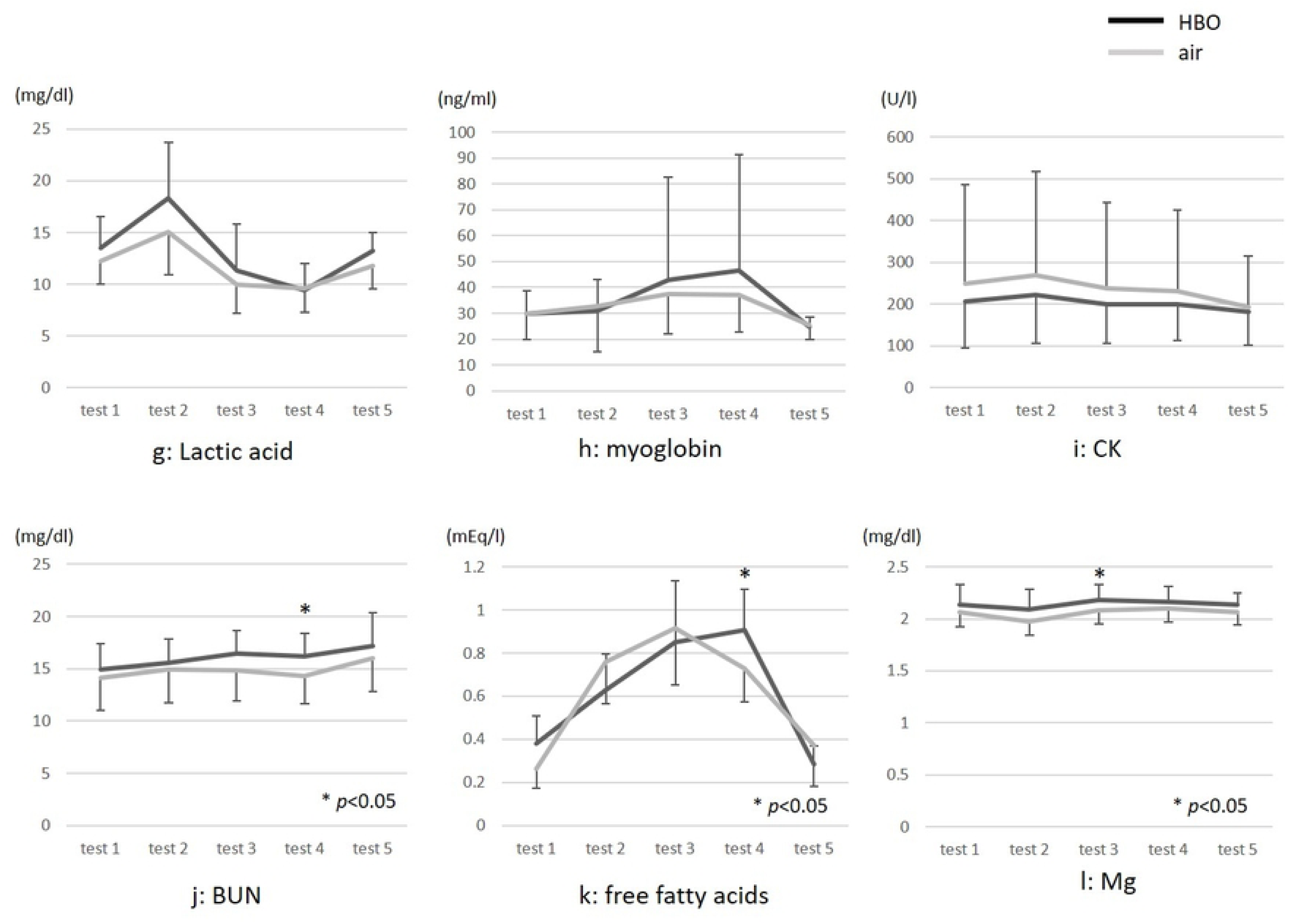
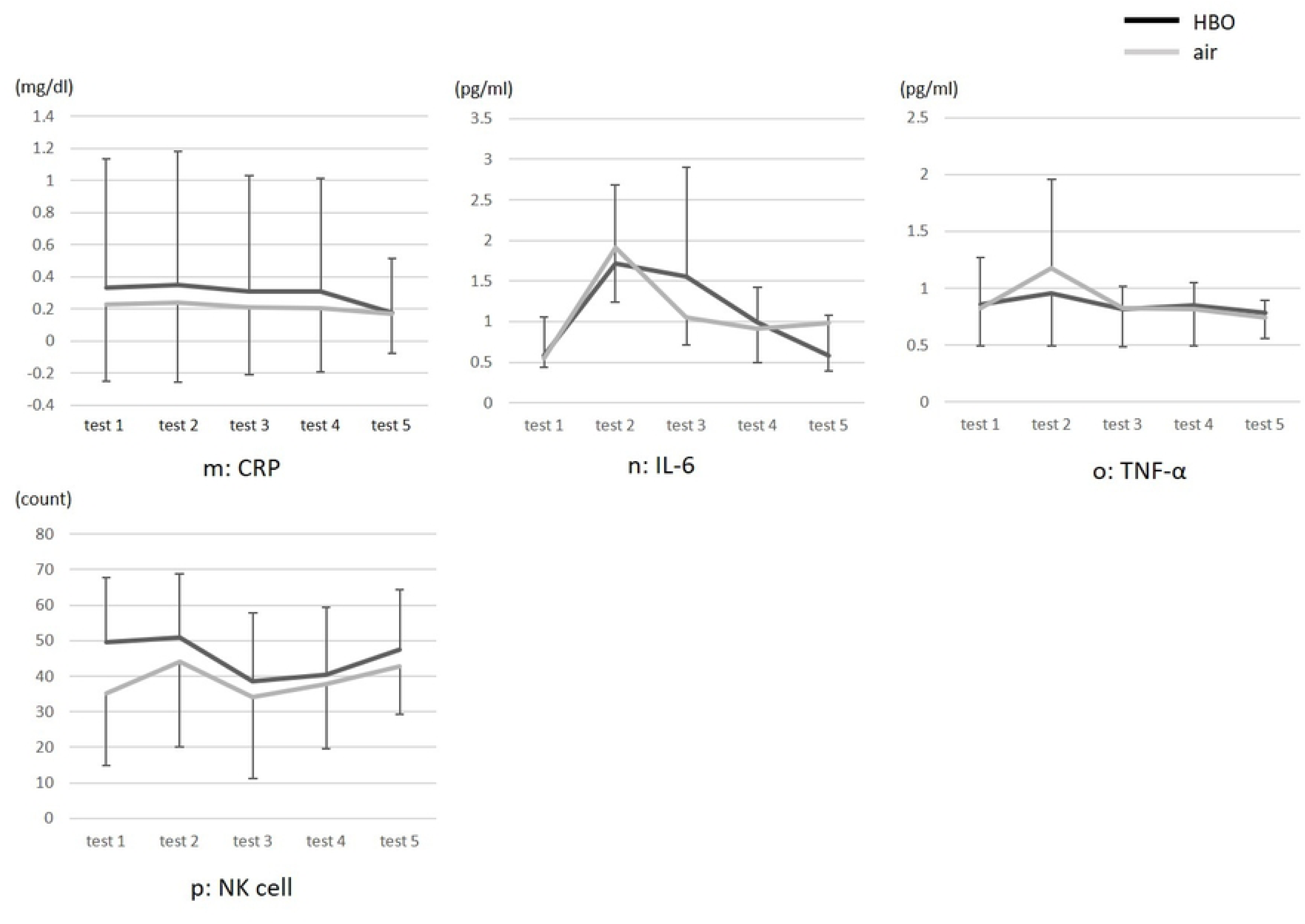
-a-p. Blood test results. CK, creatinine kinase; CRP, C-reactive protein; IL, interleukin; Mg, magnesium; NK, natural killer cells; TNFα, tumor necrosis alpha; WBC, white blood cells.

Test results indicating comparatively large post-exercise changes in both groups were particularly found for WBC, CD4-type lymphocyte, lactic acid, free fatty acids, and IL-6 levels. WBC count increased post-exercise in both groups but returned to pre-exercise levels on the following day. Neutrophil count increased and Lymphocyte count decreased in test 3, but there was no significant difference between the groups.

In the HBO group, the CD4/CD8 ratio was significantly higher in test 3 than in test 2. Although lactic acid levels markedly increased post-exercise, there was no significant difference at any time point between the groups. On the comparison of myoglobin levels for tests 2 and 3, no significant difference was found in the HBO group, and there was no significant difference at any time point between the groups.

## Discussion

### Study design and establishing the fatigue-induced workout

This study was a well-designed, single-blind, crossover randomized trial, given the high reliability of this blinded intervention, with a response accuracy rate of 53.3% and an expected value of 50.0%.

In the review of studies on the effects of HBO on recovery from fatigue after exercise, only two studies were conducted as crossover, and there were no randomized studies (23). In addition, establishing a protocol for a control group’s use of a hyperbaric chamber is generally difficult. At the very least, all groups need to be inside the chamber for the same amount of time. In this study, similar to the approach of Babul et al., the controls (air group) breathed air (21% oxygen) through a mask at a pressure of 1.2 ATA (2). According to a review of control group protocols in single-blind HBO treatment studies, rather than using the same pressure as in the HBO treatment, it is more valid to have the controls breathe air consisting of 21% oxygen at a lower ATA than for HBO (24). In the present study, when subjects were asked whether they had been breathing HBO or air at 1.2 ATA, the correct response rate was 53.3%. Approximately 50% were mistaken, suggesting that the protocol for the control group was valid. HBO studies have used various types of workouts to induce fatigue. A recent study using jiu-jitsu training sessions set workout time and intensity at 1.5 hours and the highest possible intensity (9). While some previous studies have used arm exercises (19), in this study, because a workout was needed that would affect the whole-body rather than only specific muscles, we decided to have the subjects train for a long time at a moderate intensity until exhaustion using a high-intensity training ergometer exercise bike.

Changes in lymphocyte counts can be used as an indicator of exercise intensity because the count increases until the subject finishes exercising, after which it rapidly decreases (7). Similarly, in this study, in both the intervention and control groups, increased lymphocyte counts were confirmed immediately after exercise completion, and about 2 hours later, the counts markedly decreased. As there were no significant differences in these counts between the intervention and control groups, it can be assumed that subjects in both groups had exercised with similar high intensity, and/or that the intervention had little effect.

### The Effect of HBO treatment on VAS scores for whole-body fatigue and leg fatigue

With regard to fatigue-related VAS scores, in this study, although there was significant pre- to post-intervention improvement in whole-body fatigue in the HBO group, no such difference was found in the air group. In addition, comparison of the degree of improvement between the groups showed that improvement was significantly greater in the HBO group. Leg fatigue scores in this group significantly improved between post-exercise test 2 and post-intervention test 3 from 49.9 ± 18.4 to 26.1 ± 16.9 (p<0.001) as did the scores for the air group from 54.7 ± 17.1 to 43.6 ± 24.0 (p<0.05). However, improvement was greater in the HBO group.

Akarsu et al. documented the efficacy of HBO on chronic fatigue syndrome, evaluated with the VAS, Fatigue Severity Scale, and Fatigue Quality of Life Score and found that HBO decreased the severity of symptoms and increased the quality of life (25). In a study of the effectiveness of HBO treatment for acute ankle sprain, Yagishita et al. found that VAS scores for pain decreased from pre- to post-intervention (26).

In a study on the effects of HBO on exercise-induced fatigue, Shimoda et al. documented the effects of HBO on muscle fatigue test from 50-time isometric repetitive exercises of ankle planter flexion, reporting that HBO inhibited the reduction of muscle power during repetitive exercise (27).

On the contrary, a study that measured grip contractile force in subjects in a hyperbaric chamber found that while contractile force increased with HBO, decline in force production was faster, compared with the control group, as muscle fatigue increased (28). Further, other studies have concluded that exposure to HBO before exercise does not result in improved performance (13, 21). In addition, according to the Cochrane Review, data from trials of HBO treatment for DOMS showed higher pain at 48 and 72 h after treatment (5).

In the present study, no significant differences in exercise load were found between the HBO and air groups. However, the results of VAS scores for leg fatigue after the intervention were significantly lower in the HBO group than in the air group. This suggests that HBO treatment after moderate-intensity exercise may have an effect on brain or skeletal muscles.

### Blood test results

Biochemical testing for CK serum level may have a role in monitoring healthy muscle response to training (8). In this study, although CK levels increased by about 10% in both the intervention and placebo groups after the workout, because testing for CK isozymes was not conducted, it was impossible to determine whether CK had originated from skeletal muscles. Myoglobin levels peak immediately after exercise when training has been so intense that it causes exercise-induced muscle damage and returns to its pre-exercise level within 24 h (29). As such, it is also a potential muscle damage marker (18). In our results, although no increase in myoglobin levels was found post-exercise in test 2, increased levels were found in test 3, but there was no significant difference between the groups.

Several previous studies have suggested that HBO has effects on immune system responses. Studies have shown that HBO may inhibit immune system responses (30, 31). Hou et al. investigated the effects on TNF-α and IL-6 in patients with brain tumor and reported that serum TNF-α and IL-6 levels were significantly lower after HBO than those in the control group (30). Oyaizu et al. reported that HBO suppressed the elevation of circulating macrophages in the acute phase and then accelerated macrophage invasion into the contused muscle (31). As a result, we looked at measures for numerous markers of immune system activity. However, we found no significant differences between the HBO and air groups in markers of immune system activity, including neutrophils, lymphocytes, TNF-α and IL-6.

Oyaizu et al. found that in rat skeletal muscle injured model treated with HBO, at 6 h and 24 h after injury, neutrophil markers had decreased, and at 3, 5, and 7 days after injury, macrophage movement into injured tissue was accelerated (31). While this may suggest that exposure to HBO accelerates a shift in the immune system through the early mobilization of macrophages, variation in data values was possible because of measurement timing. Other recent studies have suggested that HBO mobilizes stem cells (32, 33), and exposure to HBO after moderate-intensity training may function protectively by mobilizing stem cells to damaged muscle.

In this study, the mechanism by which HBO affects the immune system could not be determined. Further detailed studies are needed to understand these mechanisms.

### Study limitations

The limitations of this study include the small sample size, the limited number of biomarkers used, and the fact that measurements were only performed 1.5 h and 24 h after the intervention. If the measurement is performed at a different timing, the results may differ. In addition, this study only considered moderate exercise intensity. Future studies may need to evaluate the effects of HBO treatment after other types of intense exercises, such as short-duration, high-intensity exercise.

## Conclusion

In this study, subjects’ self-ratings indicated that HBO treatment may be effective in promoting recovery from fatigue after a long-duration, moderate-intensity exercise. However, this was not objectively supported by blood test results. We had hypothesized that HBO would reduce fatigue, but the results of this study could not determine the efficacy of HBO on exercise-induced fatigue. Consequently, at present, the use of HBO to hasten recovery from exercise-induced fatigue cannot be recommended.

However, because whole-body fatigue by subjective evaluation improved in a blinded trial, HBO may have some positive effects on recovery from exercise-induced fatigue via the central nerves system or skeletal muscles. More studies are needed to clarify the effects of HBO on fatigue.

## Data Availability

All relevant data are within coresponding authors personal computer.

## Acknowledgements

We thank Dr. Mitsuhiro Enomoto, Dr. Toshiyuki Ohara, Mr. Manabu Shimoda for helping in the measurement and collection of subject data.

## Author Contributions

Conceived and designed the experiment: KY, Performed the experiments: KY JA HK TO SO TK, Analyzed the data: KY HK TH TK, Wrote the paper: KY TK.

